# From Polio to COVID-19: Factors Sustaining Community Influencer Motivation in the CORE Group Partners Project’s Social Mobilization Initiatives for Vaccination in India

**DOI:** 10.64898/2026.07.22.26358734

**Authors:** Anna Schurmann, Arup Kumar Das, Sanjna Sinha, Pankaj Mishra, Balkrishna Yadav, Parul Ratna, Jitendra Awale, Manojkumar Choudhary, Kathy Vassos Stamidis, Hibret A. Tilahun, Henry Perry

**Affiliations:** Department of Global Health and Development, Faculty of Public Health and Policy, London School of Hygiene and Tropical Medicine; Uttar Pradesh State Transformation Commission, Health Cell; Independent Research Consultant; Center for Social Medicine and Community Health, Jawarhal Nehru University; Tattva Foundation; CORE Group Partners Project Secretariat, India; CORE Group Partners Project, Washington, DC. USA; Johns Hopkins University

**Keywords:** Polio, vaccines, community health volunteers, motivation, India

## Abstract

**Introduction:** Engaging Community Influencers (CIs) was a key strategy for polio eradication in India, also contributing to routine vaccination and COVID-19 vaccination. CIs were high status individuals selected for their pre-existing reach into vaccine-hesitant communities. This paper describes the factors that kept the CIs engaged over time. The findings add to our understanding of what drives CHW motivation.

**Methods:** We performed a thematic analysis of 65 in-depth interviews conducted across six study districts of Uttar Pradesh, involving 36 CIs, 18 CGPP field staff, nine project managers and two representatives from other stakeholders.

**Findings:** The most important motivational factors that emerged from the research were improved social status, altruism, and effective working relationships. The inputs required to foster these were information support; providing respect and visibility; and fostering robust working relationships between program staff, CIs, CHWs, and government functionaries.

**Conclusions:** CIs’ engagement in the program maintained and bolstered their pre-existing status. Deploying and sustaining this cadre requires a strategic approach to maintaining motivation over time. Providing respect and visibility and maintaining effective working relationships is key to sustaining the CIs’ motivation.

**Key Messages:**

- The Community Influencer approach, initially introduced for polio eradication, has evolved over time into a vital strategy for increasing routine immunization and COVID-19 vaccination coverage.
- Community Influencers are high status individuals, selected by program (CGPP) staff for their pre-existing status in vaccine-hesitant communities.
- This study looks at how the motivation and engagement for this cadre was sustained over a 20-year period.
- Several key factors sustained the Community Influencers’ engagement over time, including a pre-existing commitment to their community, the program successes, visibility and respect; the working relationships with program staff and government functionaries; and, finally, the Influencers’ status within the community.
- Considering that many health systems struggle with a demotivated workforce, these findings provide practical guidance for fostering health worker engagement.

## Introduction

Community health workers (CHWs) are considered key to achieving Universal Health Coverage and Health for All.(1) These cadres can extend service to vulnerable and excluded populations, provide care and messaging in a culturally appropriate manner, improve access to services, address inequities in health status, and improve health system performance and efficiency. Globally, across many different national and regional program CHW cadres, a variety of factors engage and motivate CHWs, including financial, social, and intrinsic incentives. However, maintaining motivation often poses a significant challenge to performance, scalability and long-term sustainability of CHW programs.(2–4)

Community influencers (CIs), a cadre of community health volunteers, are respected members of their communities who help build trust to facilitate interactions between the health system and traditionally neglected and underserved populations. CIs were selected for their pre-existing reach within vaccine-hesitant communities. CORE Group Partners Project (CGPP) staff mapped communities and identified vaccine-resistant families based on the nature of their resistance, and suitable CIs were identified and briefed to persuade people for vaccination. The CGPPs CIs have now been active for up to 20 years, first working specifically for polio vaccination, then on routine immunization more broadly, and more recently on COVID-19 vaccination drives. CIs and *the Community Influencer approach* now have an established record of building vaccine confidence and reducing inequalities in vaccine outreach and coverage.(5)

### Project Background

The CGPP, a consortium of international and national non-governmental organizations (NGOs), funded by the United States Agency for International Development (USAID) has been working to eliminate polio across several countries since 1999. The CGPP operations in India, which concluded in September 2024, were implemented by three international NGOs - Catholic Relief Services (CRS), Adventist Development and Relief Agency (ADRA) and Project Concern International (PCI) – along with six local NGO partners. These organizations worked across 12 districts of Uttar Pradesh, two districts of Assam and two districts of Haryana to support CGPP’s goal of polio eradication through mobilizing communities for polio and other routine vaccinations and maintaining population immunity. CGPP India’s direct intervention in Uttar Pradesh districts has reached approximately 3.8 million people.(6) In India, the CGPP drove reductions in polio vaccination refusal which, in part, was due to the strong links created with CIs.(5,7) The CGPP had several cadres at different administrative levels in the field, including District Mobilization Coordinators (DMCs), Block Mobilization Coordinators (BMCs) and Community Mobilization Coordinators (CMCs).

The CGPP adopted a sustained, multi-pronged, community-focused strategy over a prolonged period to counter vaccine hesitancy. Its staff (CMCs and BMCs) mapped all households and performed door to door activities to identify and enlist under five children those were eligible for vaccination and encouraged households to get vaccinated.(8) CGPP staff (CMCs and BMCs) supported government vaccinators in disseminating information and setting up immunization booths. In addition, it conducted various social mobilization activities, such as, street plays and rallies wherever possible to spread information about the immunization programs.

In 2014, once polio was eradicated from India, as part of the national ‘endgame’ strategy, the key learning and promising practices from polio were applied in government routine immunization programs to augment coverage. Under the banner of social mobilization, one of the promising practices that were identified and applied under CGPP in the context of Routine Immunization was engaging CIs to mobilize communities, especially in overcoming community resistance to getting children fully vaccinated. CGPP’s use of CIs spanned three different immunisation program domains: the polio program, routine immunization of pregnant women and children, and most recently the COVID-19 immunization program.

While the work of the CIs has been perceived anecdotally to be impactful, the extent of their role has not been captured comprehensively in existing literature in India or globally.(9) There has been little focus on the motivational factors that kept them engaged over time. While recruitment and deployment of CIs was ongoing, many remained engaged for prolonged periods of time (unfortunately data is not available on the average length of tenure). More research is needed to understand how CIs are motivated to support immunization campaigns and the barriers and facilitators of their engagement. This will, in turn, provide guidance for the future use of CIs to support outreach activities in India and globally. The overall aim of this research is to identify factors that contributed to the motivation, engagement, and length of tenure of CIs in successfully increasing childhood immunization (including polio) and COVID vaccination rates.

## Methodology

To support this aim, we adopted the following objectives:

- Identify the strategic approach and inputs from CGPP in terms of working with CIs
- Understand the motivation of the CIs
- Identify the challenges and enabling factors that helped CIs address vaccine hesitancy among families.

### Theoretical Framing: Theories of Motivation and Behaviour Change

To develop a theoretical framework to guide the study, we reviewed the literature around motivation factors for CIs and CHWs and developed a provisional framework. We then refined this through analysing CGPP’s secondary data and reviewing primary qualitative data.

To understand the motivation of CIs, we explored theories around behaviour change and motivation. We looked at the empirical and theoretical literature around community health worker (CHW) motivation from both India(10–15) and globally, including self-determination theory and concepts such as social capital (2,3,16) (17–20) (21–23) The research team also reviewed empirical work around health worker motivation more generally(24)(25).

Much of the empirical work around CHW motivation focuses on programmatic incentives ^4^, workplace conditions(13,15) (extrinsic factors), or individual-level cognitive motivational factors(14)(12) (intrinsic factors). There is less consideration of the social, relational, and community-level factors that might motivate a socially anchored volunteer cadre.^3^ Social identity theory was incorporated to address this gap in prior work around community health worker motivation.(16) Social identity theory considers both individual and contextual factors to understand how individual cognition depends upon interpersonal relationships and group membership. It has increasingly been used around questions of motivation, including CHW motivation.(16,26) It helped form our hypotheses; that the CIs were motivated by their commitment to the shared identity that is their “community”, their *status* within it, and the *status* of the community itself. (17,19) In this context *individual status* includes (but is not limited to) socio-economics status, class, caste, resource-holding potential and most importantly for this study, social influence. It refers to the influence one has over group decisions and resource distribution. The *status of the community* refers to the level of health and wellbeing in a relative sense (compared over time or between different communities). These hypotheses were used to create a provisional “conceptual model” that was tested, refined, and recreated throughout the research process. This conceptual model guided data collection and analysis and eventually came to reflect the research findings. This is depicted in Figure 1, which illustrates the identified status benefits in the top row, motivational factors in the middle row, and in the bottom row, the program inputs and activities, describing stepwise how the program supported motivation over time, like a theory of change model.(27)

**Figure 1:**
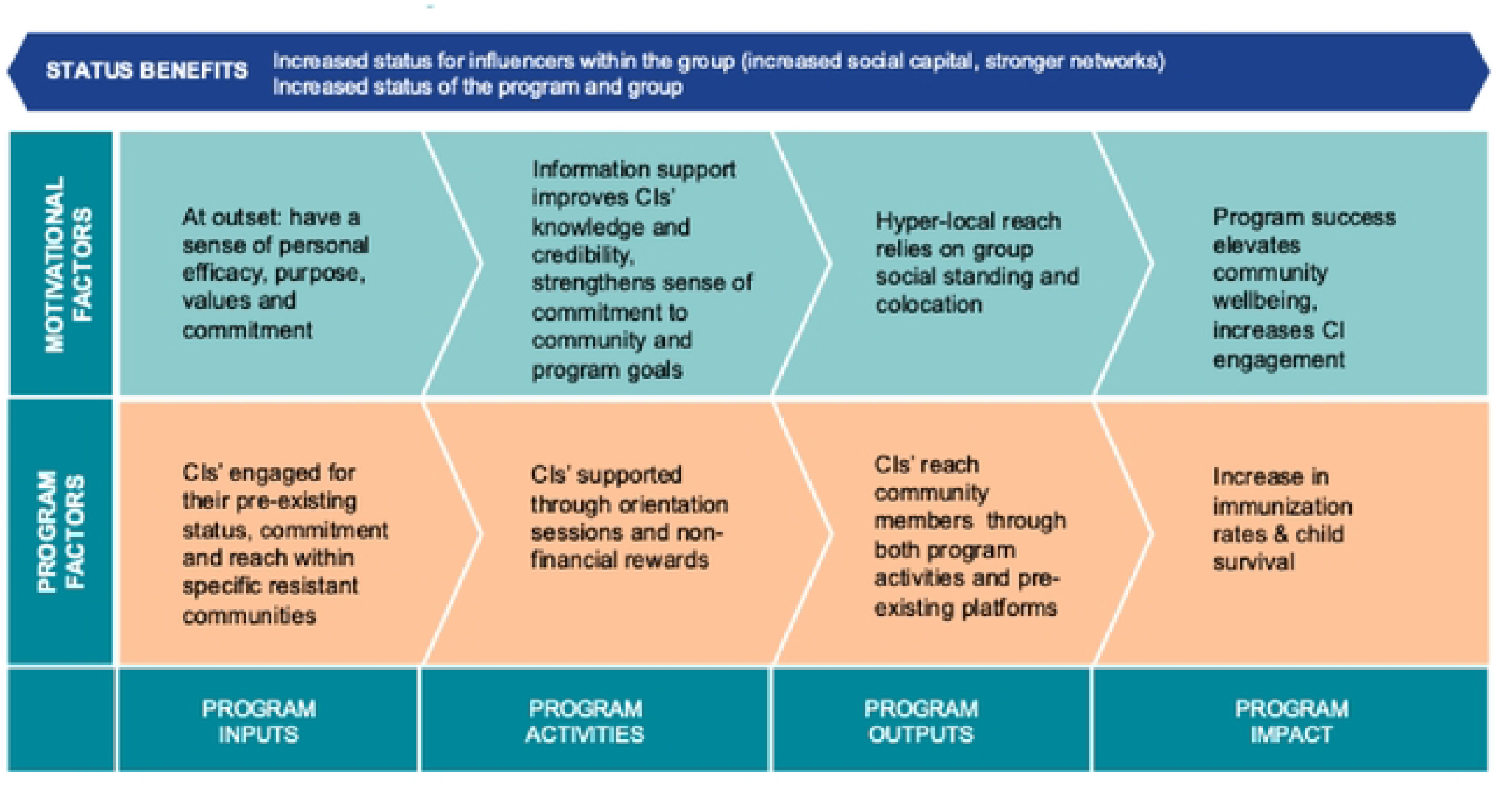
Conceptual Model of Factors Influencing the Engagement of CIs

### Study Population

CIs are typically high-status individuals within their community performing influential and highly interconnected roles. Data from project records in the six sample districts indicate that most of the CIs were male (86%), married (96%), and Muslim (52%) (versus 19% in the general population). The median age of the CIs was 42 years, with 50% of them having completed higher secondary education. Common (primary) job roles for the CIs included village councillors (Panchayat Raj members) (12%); teachers and other government workers (11%); local medical practitioners (9%); ration shop owners (8%); and religious leaders (6%). *A ration shop (or fair price shop) sells grain, sugar and oil at subsidized rates, as part of a food security scheme under the Public Distribution System (PDS), run by state governments within India. Some ration shops are run by individuals, others are run by panchayats, self help groups or co-operative societies*.

In-depth interviews were conducted with selected CGPP staff and CIs in the selected six districts. The sample was purposively selected through liaison with district-based CGPP staff and an actor mapping exercise. It consisted of stakeholders representing project roles and capabilities as well as CIs spread across all six districts. The CI sample profile aligns with the general profile of community influencers. The completed sample consisted of a total of 65 interviews, including 36 CIs,18 program staff, 3 CGPP secretariat members, 6 members of CGPP Private Voluntary Organizations, 1 WHO official and 1 District Immunization Officer.

Interviews were conducted in late 2022 and early 2023 by a primary interviewer with the support of a notetaker. All participants were provided with protective measures against COVID-19; the interviewers and the notetakers were provided with N95 masks, sanitizers, and shields. Before starting the interview, it was mandatory to wear masks and maintain proper physical distancing during participant contact.

Regular debriefing sessions were held with the data collection team and the principal investigators to identify preliminary key themes and discuss any field-based challenges. All interviews were recorded, then transcribed and translated into English. They were then quality-checked for clarity before being uploaded for analysis with NVivo.(28) The analysis was conducted thematically according to the conceptual framework and then summarized in narrative format. Emergent themes were also documented through memos and annotations, and these were also summarized.

### Research Ethics

All interviews were conducted only after obtaining informed consent and taking all precautions regarding confidentiality and data protection. Institutional Review Board (IRB) ethics approval was received from Sigma Research and Consulting, Pvt Ltd (10064/IRB22-23).

### Findings

The findings cover several pre-identified and emergent themes, with the intent to tell the story of the CIs as well as explain the factors that maintained their engagement over time. First of all, vaccine hesitancy is the problem the CIs were engaged to solve, and it is important to understand it in its local context for polio, routine vaccination, and COVID-19 and across different communities. This is described in the first instance. The profile of the CIs is important for their effectiveness and engagement over time, so the next sections describe the process through which they were identified and engaged, and the perceived characteristics of an effective CI. Subsequently, the key activities and project support provided to the CIs is described. The concluding section is the key focus of the paper: the factors that fostered motivation and engagement over time.

### Vaccine Hesitancy

The CIs were deployed to overcome challenges around vaccine hesitancy. Resistance to vaccinations in Uttar Pradesh took several overlapping forms and changed over time. Initially, resistance to polio vaccination was concentrated in specific minority religious communities and based on mistrust of the government. Participants recalled that there were rumours and misinformation circulating that the virus in the vaccines was grown on a substrate of monkey cells or included pig cells and so would be prohibited under Islam and Hinduism. The initial messaging emphasizing “two drops” for polio vaccination, unfortunately, recalled earlier messaging from a coercive family planning program that emphasized two children per family. This meant there was a fear that the polio drops would make the recipient infertile. Later, resistance to childhood immunization (including polio immunization) was more diffuse and based on a complaint that the government’s development priorities should be based on locally tangible needs, such as food rations, electricity, and roads, rather than polio immunization.

Participants shared several colourful stories about active community resistance to the polio vaccination program. One older woman rolled her granddaughter in a rug to hide her from vaccinators. Another woman threw her baby out of reach of the vaccinators (in both cases, the children were unharmed). Staff reported having boiling water or dirty drain water thrown at them in narrow lanes where it was hard to escape. One vaccination team recalled being treated with kind hospitality, taken to a room and given drinks, then being locked in for the rest of the day.

*In 2008 in [a particular area] they had very narrow lanes where they would throw boiling water on us when we entered. I still have the burn wounds*.

A CGPP India Staff Person

### Recruitment of Community Influencers

The elevated level of resistance to vaccines hampered vaccine outreach and highlighted a lack of trust between the health system and communities. CGPP staff sought to overcome initial program limitations by co-opting local community leaders as CIs to increase impact, especially among resistant households. As trusted members of the community themselves, CIs could help spread appropriate information, counter rumors, and alleviate fears through everyday interactions instead of through the formal channels of communication available to program staff.

The success and enduring motivation of the CIs depended on identifying and deploying the right people. CIs were selected because of their existing status, reach, and influence.

When commencing house-to-house outreach in a particular geographic area, program staff mapped potential CIs through first mapping the local communities and sites of vaccine resistance (a process called *microplanning*(8)). These maps were routinely updated at the time of house-to-house vaccination drives. Staff would then interact with community members to identify individuals with existing influence in the community. They also approached community leaders such as the elected village panchayat head (*Pradhan*) or religious leaders like the local priest or imam to recruit them as allies of the program. As one CMC explained, “Everywhere there is one person to whom everyone listens, so we meet him and take support.” CIs could be ration-shop owners, teachers, informal healthcare providers, or members of local associations such as the weavers’ association. In addition to the strategic efforts of the program staff to identify those with pre-existing influence, CIs were also identified opportunistically through the course of routine advocacy among communities.

CIs were selected because of their existing status, reach and influence, not their opinions or existing knowledge. These individuals might have initially been advocating against vaccination but once recruited, briefed, and convinced about the project, would provide active support.

To recruit CIs and foster their commitment, CI involvement was presented as a form of social service to their respective communities. The program staff also suggested that they would achieve respect and status from working with the project, with potential to expand the reach of their business or political support base. The findings indicate these suggestions were realized, to varying extents.

The CIs’ reach and impact was typically locally specific. However, this varied across CI types. Religious leaders were typically restricted to their congregation or community. Others, such as political leaders, doctors, and teachers, had a broader reach. Study participants also reported that the CIs’ reach expanded over time. Multiple CIs were usually engaged in the same community to ensure that the same message was spread and reinforced in diverse ways.

These processes of convincing individuals to voluntarily support the program were thus iterative and ongoing. There was not a consistent approach in terms of number of CIs per community, or level of contact with each household. Instead, program staff responded to the specific nature of vaccine resistance in each community by recruiting CIs as appropriate. DMCs, BMCs, and CMCs worked continuously to persuade, clarify, and reinforce the commitment of CIs. They used multiple strategies to ensure ongoing engagement of the CIs by sharing up-to-date information with them. A DMC emphasized that regular interactions between program staff and the CIs were key:

*We conducted meetings every month. We kept stirring their (CIs’) minds. If you have a friend and don’t meet him for quite some time, how will you maintain the friendship? Regularity is necessary*.

A CGPP India Staff Person

### Characteristics of an Effective CI

If the success of the program relied on selecting the right people, then what did the “right person” look like? The characteristics of an effective CI varied according to the community and the reason for vaccine refusal. Sometimes, the right person was a religious leader, other times, a ration dealer, a doctor, or a pharmacist. The first step was to understand the reason for refusal and then match the CI accordingly. In more specific terms, reported attributes of an effective CI included speaking well, being educated, having a pre-existing inclination towards volunteerism and social service, having good standing and respect in the community and being a good listener (which was especially reported by female CIs). CIs described their social position through recounting their acts of service, helping fund weddings of poor young women, being a source of advice, and being called upon to settle disagreements. Several CIs also mentioned the support of their family as key to their effective engagement with the program. One program staff member describes an effective CI’s abilities as being able to move people; “he used to say things in a better way than we used to tell him, and it would touch peoples’ hearts…”

*The people understand the way I speak, when a government officer comes, he will talk in his way, but I talk in a familiar way with people, so whatever the public will tell us, then they will not be able to say that to the officer. They share their problems with us, and we talk to them in their language, and in that language only we make them understand how it will happen…if someone comes from outside then it is different*

A Community Influencer

From the perspective of program staff study participants, one of the most important attributes of a CI was their availability. CIs contributed to the program according to their availability as they balanced work, family, and community commitments. Varying availability and capacity were something the program staff were comfortable adapting to, even though it was sometimes challenging: “Someone is small or big, as per their capability they have been giving their contribution.” CIs typically had flexible schedules, and most did not report any disruption to their livelihoods due to their work as a CI. For some CIs, their businesses expanded as a perceived result of their involvement, through increased status and visibility.

From the perspective of the study participants themselves, CIs took pride in their skills and influence within the community and were willing to work hard to maintain this stature. They considered themselves as being trustworthy, of good character, knowledgeable, and respected. These individuals were inclined to offer guidance and advice to friends and family on various matters even before being recruited by the CGPP. One CI attributed her effectiveness as a CI to coming from a prominent and well-known family:

*…everyone in my family is educated. So, people from the community respect my family and me already… I am a known face in the locality and receive respect in the Madrasa as well. They listen to me*.

A Community Influencer

### Community Influencer Activities

Interpersonal communication with individuals and families to persuade them to get vaccinated, especially in the case of vaccine resistant individuals, is a primary role for the CIs, and key to the success of the entire CGPP, as this quote illustrates:

*Communication is really important. Things will happen only when we talk to people about it. By talking we not only tell them what we want but we also get to know what they think about a certain thing. Then only we will be able to bring about the change we want*.

A Community Influencer

CIs used different tactics with different people based on their own unique situations. They used parables, quoted religious texts, cited firsthand experiences, and used communication materials to pass on messages. CIs would often help each other or ask other influential members of the community to help convince resistant individuals. During the COVID-19 pandemic, CIs demonstrated their commitment to their communities by proactively helping arrange water and soaps for families, distributing masks and sanitisers, promoting maintenance of social distancing, and enforcing government guidelines.

One CI described himself as a “guarantor” who has taken responsibility for the safety and effectiveness of the immunization program to build trust between the community and the health system. This included offering to pay for any follow up care required as a result of immunization or accompanying families to the doctor or hospital in the event of an adverse outcome. He said, “I tell them that **I am there** so please take the polio drops.”

Program staff actively sought to involve CIs in their activities. For example, CIs facilitated meetings of various target groups such as mothers, adolescents, and fathers to overcome vaccine resistance as well as raise the profile of the CIs themselves, as one DMC explains:

*We have many kinds of group meetings such as Mata Baithak [mothers’ meeting] or Saheli meetings [adolescent girls’ meetings]. We would make sure the influencers attended them. When they attended these meetings, they felt honoured. We make it a point to invite them… All the people we want to target are present there and included the influencer too, for better impact*.

A CGPP Staff Person

### Programmatic Support for Community Influencers

CIs were selected because of their existing reach, not their opinions or existing knowledge. The program staff supported CIs at each step to ensure their effectiveness. The CIs were briefed both formally through scheduled monthly meetings (which included a training component) as well as informally through interactions with program staff and government frontline workers on various aspects associated with the diseases, vaccines, and relevant health services. CGPP BMCs and CMCs would provide content such as pamphlets, banners and communication materials that were used by the CIs to mobilise community members. The accuracy of the information provided through the program ensured that the CIs were seen as credible in the eyes of the community, as a CI explained:

*Whatever information we provided … we were proved not to be liars. For instance, we made people understand something and then if we were not able to hold our ground on that information, then people’s trust would break. When the organization proved to be credible, they supported us…and that is the reason we have been trustworthy until now*.

A Community Influencer

Over the years of engagement, program staff and CIs became close, with one participant describing their relationship as being like family. The strong interpersonal relationships that sustained the program were clear across all participant types. The CIs appreciated the transparent and egalitarian communication between the program staff and the CIs.

Some CIs downplayed the programmatic support provided to them; they presented themselves as largely self-sufficient and able to solve their own community’s problems without outside help. These CIs described the need for minimal program inputs in the form of information, coordination, and the backing of a program structure to maintain their work. Nevertheless, there was a broad acknowledgment that they could not continue their work without some kind of programmatic support.

### Motivation and Enduring Commitment

“I have gained a lot of self-confidence when I see that people are listening to me and are following my advice. I feel very satisfied when I see that people are being influenced by me and what I say.”

A Community Influencer

There were multiple motivations for people to initially join as CIs: commitment to the community, religious motivation, a disability in the family, and self interest in terms of expanding their business or building a political base. Several CIs described a family history of polio among siblings, children, and childhood friends with the disease, all inspiring CIs to commit to this work. The quote below is an example:

*Till 5th class, we studied together, and we used to carry him as the roads were not good and there was sewage in our village. There are lots of people who have suffered from polio in our village. Some use crutches, and some use their hands to walk*.

A Community Influencer

Another CI reported being able to win the Panchayat election due to the goodwill generated through his work as a CI. One CI described how in the context of ongoing community disputes, he wanted to shore up power and influence as a protective factor:

*Around 2007, I was constructing my house, and a person from the [land-owning] community created a lot of disputes. That was the time when I decided that I needed some kind of political support, so I became active in politics and became a social worker*.

A Community Influencer

CIs were not compensated for their efforts financially. Indeed, the voluntary nature of their work added to their perceived legitimacy within the community – while program staff were doubted because they were paid to spread a particular message. Altruism can be seen as a source of both motivation and legitimacy. One CI said: “I am not telling anything by making up anything. Everything is genuine.”

One of the ways altruism manifests itself is the recognition that collective community well-being promotes well-being at the individual or the household level. This is described by one CI as follows,

*I have learned that by helping people, you can stay happy. If you are happy, then other people will be happy with you and you will be motivated to live life…If someone is unhappy and you are happy in your house and eating cashews and raisins and other people sleep on an empty stomach, then eating cashews and raisins was of no use*.

A Community Influencer

Altruism is also key to how CIs described their communities, in idyllic-sounding terms, as places where everyone is loved and cared for. It also relates to how they described themselves as exemplars of selflessness and leaders in their communities through their acts of service and beneficence.

Aside from community well-being, motivation was also described in terms of working relationships and shared challenges. This created robust and fond relationships between CIs and program staff, with several study participants describing the relationships as like family (as mentioned earlier). One CI described the increased enjoyment he experienced after overcoming early challenges;

*Since we got connected to the CMC, we faced a lot of problems. After going through all of that, we now enjoy our work more. We have tolerated so much. At least now people are happy. We do that work now and we like it*.

A Community Influencer

While they were selected for their existing reach and profile, the CIs’ role in the program was reported as becoming part of their identity in the community. The success in eliminating polio became the CIs’ success. This manifested itself tangibly through expanded political platforms, more profitable businesses, and more visibility in the community: “There is no monetary return, but people know us and this itself is enough.” CIs also describe their own increased authority, with families much more willing to listen to them now.

“There are two things: one is satisfaction, and one is respect…We have seen that respect is a diamond, a man’s achievement. Nothing has its equal. The person who understands what this is, the one who enjoys it, never leaves it.”

A Community Influencer

Program staff were cognizant of how respect was able to maintain CI engagement and leveraged this through giving CIs visibility at events and meetings (sometimes at 5-star hotels or other high-status venues) where they were felicitated by senior government officials and applauded by their peers, thereby raising their stature within the community. Recognition and honours were sometimes amplified through social media and the local press. The quotes below demonstrate this from both sides: from the program staff member, and from the CI.

*We tried to recognize their efforts…in various platforms. It was easy for us to call them and give them a chair at the district task force meeting. For people from a remote village, direct connection with the district magistrate was a big thing. The district magistrate is saying their name. It was a big honour that the district magistrate knew them*.

A CGPP India Staff Person

*At that time, a certificate was given to me for the work I had done during the Covid pandemic. I have been treated as a special person. That time, I felt that I had done good work, and I was getting honoured for that. My photo was circulated in social media and people told me, ‘You deserve this, and you had the right to be honoured’*

A Community Influencer

Giving respect and recognition for the CIs, was a key program strategy for fostering engagement. The CIs had the respect of the community, and for that reason, they had the respect of the project. As a CGPP secretariat member said in the following quote.;

That is the main thing. You respect them. You know that he’s not a trained MBBS doctor… but he has a big following of clients, so he’s obviously doing something right…He’s getting the respect of the people. We give him the respect as well, and we teach him also.

### A CGPP India Secretariat Member

CIs also received certain privileges, either indirectly by virtue of their proximity to government officials or directly through their association with the vaccination program. For example, CIs were prioritized to receive the COVID vaccinations during the initial phase when there was a scarcity, CIs were also prioritized to get visas and subsidized flights for *haj*. *The Muslim pilgrimage to Mecca in Saudi Arabia, held annually in the last month of the year. All Muslims are expected to go once in their lifetime if they can afford it.* One program staff person also saw proximity to the government as a motivating factor in terms of the perceived protection it provided:

*They think by supporting the government they can save themselves a little from the pressure that the government puts*.

A CGPP India Staff Person

Program success did not just affect the motivation of the CIs. It improved community engagement more generally. One program staff member described initial challenges in mobilization, with enthusiasm and momentum increasing over time:

*Previously, if we invited 40 people to community meetings only 20 would appear, and the rest would think of it as not so important. Nowadays, when we organize community meetings, we see a crowd of people voluntarily joining the meeting. People now know about the importance of immunization and want to know more*.

A CGPP India Staff Person

## Discussion

The focus of this study was to identify motivational factors for CIs, and the programmatic inputs required to foster this. While multiple motivational factors emerged from the research, the three key factors were relational and prosocial: *altruism, status and respect*, and *working relationships*.

### Altruism

Altruism is defined by as “a moral norm [which] implies certain social expectations of helping others in different social contexts.” This suggests that altruism is more than helping others, it also reflects social expectations: you help the collective.(29,30) We find this definition useful for this setting. While there were several reasons that prompted CIs’ initial engagement, CIs were typically selected because they were already leaders within their community. In their self-image as leaders, they described themselves in terms that reflected the priority given to social and moral values; altruism, selflessness, and a commitment to ‘social work.’ In this context, altruism is a source of legitimacy, with CIs and program staff reporting that the CIs are more likely to be trusted and believed in their communities because they are not being paid for their efforts; they are acting out of a commitment to the community’s well-being. This is also reflected in how they described their communities - as places where people support and care for each other. Well-being and happiness are also described as collective rather than individual attributes, in that a person can only achieve well-being and happiness if everyone in the community has achieved them.

Research throughout the world has consistently found positive health and wellbeing outcomes from volunteering.(31–34) Furthermore, research by Aknin et al. suggests that the wellbeing impacts of volunteering are greater when they promote social connection.(35) In many ways, prosocial acts contribute to social cohesion and community resilience.(36) Our findings add to this body of knowledge – with CIs reporting feelings of peace, satisfaction, and pride at the work they have done. Thus, working altruistically (sincerely and strategically) has impacted CIs by providing them with a sense of personal satisfaction and achievement.

### Status and respect

Our research findings suggest that being part of the program augmented the status of CIs and that this played a strong role in keeping them engaged (as shown in Figure 1). This was derived from several factors: their proximity to power through the program; the program’s success and how this reflected well on them; and, relatedly, the visible decline in illness and disability. A key factor that speaks to status is respect: the respect given to the CIs by the CGPP and government functionaries and the respect they received from communities. This aligns with other research findings around CHW and health worker motivation. For example, a review of evidence of factors influencing health worker motivation found that appreciation and respect was a factor in 70% of studies. (37)(4) More specifically, research into factors motivating community health volunteers in Mozambique found that being given respect for program success was key.(38,39) This ties in with social identity theory, which suggests that people will be motivated to contribute to an effort if it improves the status of their community and of themselves.(36)

### Working relationships

Motivation was also fostered through working relationships and the shared successes and challenges associated with them. This aligns with findings from other research about health worker and community health worker (CHW) motivation, from India and globally.(37)(3,11) Our findings demonstrate that the working relationships between program staff and CIs mutually built skills and competencies – a cognitive source of motivation. These working relationships also provided a sense of being valued and supported – an emotional source of motivation.(2) We would also suggest that working relationships contributed to a sense of affiliation with each other, and the project at large, in alignment with social identity theory.(36)

Reflecting on these findings, the conceptual model we presented in Figure 1 prioritizes prosocial and relational elements of motivation. The model also maps the project inputs required to foster this motivation, including fostering robust working relationships and providing respect and visibility and ongoing informational support. These findings around relational and prosocial motivational factors give depth to an understanding of what a “people-centred health system” might look like.(40) These inputs are also within the capabilities and resources of most NGOs and government institutions. In a global context where many low- and middle-income countries’ health systems struggle with demotivated health workers and health worker shortages, these findings add to the literature on health worker motivation and community engagement. At the same time, these findings are specific to a socially anchored volunteer cadre working in an economically under-developed area with limited opportunities for social and economic advancement.(2)

### Limitations

The study has several limitations. First, there was selection bias; all the respondents were purposively selected. Nonetheless, the demographic profile of the CIs in our sample matched the profile of the larger population of CIs in the six study districts. Even so, it is possible that the selected CIs’ opinions did not match those of the larger population.

Secondly, social desirability bias may have been present. Several of the CIs saw participating in interviews as an honour and a gesture of recognition. Many positioned themselves in a kind of ambassadorial role and provided accounts of themselves and their communities which were highly positive and self-congratulatory. Including program staff in our study sample helped us achieve a more well-rounded impression.

The third limitation is recall bias. The program had been ongoing for two decades and CIs’ association with the program occurred at various times during this period, so their collective memories may have been incomplete. Despite probing, the research team struggled to get a sense of a linear narrative with dates and clear program milestones that could have contributed to a social history of the program, especially in interviews with the CIs themselves.

## Conclusions

CIs were typically individuals within the community with pre-existing high status and influence. These individuals helped create an enabling environment for the immunization program through interpersonal communication to persuade resistant individuals and families. The findings from this study support the hypothesis that the key motivating factors for the CIs was a commitment to the well-being of the community and the opportunity for improving their status within it. Additional findings identified respect and successful working relationships as additional key motivating factors, which were also a key to their effectiveness. These findings are a valuable addition to the literature on community health worker engagement but are also specific to a socially anchored volunteer cadre.

## Data Availability

The qualitative data that underpin this research cannot be made available, due to ethical conditions associated with the research. The in-depth interviews were conducted with a promise of confidentiality that they would be used for the specified research only. The study participants - Community Influencers (CIs), CGPP program staff, and other stakeholders – hold prominent positions in their communities, which would make them recognisable as a result of information they provide, even following redaction. Limited access to de-identified transcripts may be feasible, subject to approval by the Research Ethics Board at Sigma Research and Consulting Pvt Ltd (approval no. 10064/IRB22-23) by contacting: Manoj Choudhury Global Technical Advisor, MEAL CORE Group Partners Project World Vision 300 I Street NE, Washington, DC 20002

## Notes

### Competing Interest Statement

The authors have declared no competing interest.

### Author Declarations

Sigma IRB, CIN No: U74140DL2008PTC182567

